# The association between experience of COVID-19-related discrimination and psychological distress among healthcare workers for six national medical research centers in Japan

**DOI:** 10.1101/2022.08.03.22278304

**Authors:** Rachana Manandhar Shrestha, Yosuke Inoue, Shohei Yamamoto, Ami Fukunaga, Makiko Sampei, Ryo Okubo, Naho Morisaki, Norio Ohmagari, Takanori Funaki, Kazue Ishizuka, Koushi Yamaguchi, Yohei Sasaki, Kazuyoshi Takeda, Takeshi Miyama, Masayo Kojima, Takeshi Nakagawa, Kunihiro Nishimura, Soshiro Ogata, Jun Umezawa, Shiori Tanaka, Manami Inoue, Maki Konishi, Kengo Miyo, Tetsuya Mizoue

**Affiliations:** Department of Epidemiology and Prevention, Center for Clinical Sciences, National Center for Global Health and Medicine, Tokyo, Japan; Department of Health Science, Health Promotion, Nippon Sport Science University, Tokyo, Japan; Clinical Research & Education Promotion Division, National Center Hospital, National Center of Neurology and Psychiatry, Tokyo, Japan; Department of social science, National Research Institute for Child Health and Development, Tokyo, Japan; Disease Control and Prevention Center, National Center for Global Health and Medicine, Tokyo, Japan; Division of Infectious Diseases, Department of Medical Subspecialties, National Center for Child Health and Development, Tokyo, Japan; Center of Maternal-Fetal, Neonatal and Reproductive Medicine, National Center for Child Health and Development, Tokyo, Japan; National Center Hospital, National Center of Neurology and Psychiatry, Tokyo, Japan; Department of Frailty Research, Research Institute, National Center for Geriatrics and Gerontology, Aichi, Japan; Department of Social Science, Research Institute, National Center for Geriatrics and Gerontology, Aichi, Japan; Department of Preventive Medicine and Epidemiology, National Cerebral and Cardiovascular Center, Osaka, Japan; Division of Cohort Research, Institute for Cancer Control, National Cancer Center, Tokyo, Japan; Division of Prevention, Institute for Cancer Control, National Cancer Center, Tokyo, Japan; Center for Medical Informatics Intelligence, National Center for Global Health and Medicine, Tokyo, Japan

**Keywords:** COVID-19, healthcare workers, discrimination, psychological distress, mental health

## Abstract

**Background:** Discrimination has been identified as an important determinant of negative mental health outcomes. This study determined the association between the experience of COVID-19-related discrimination and psychological distress among healthcare workers (HCWs) in Japan.

**Methods:** This cross-sectional study conducted a health survey among 5,703 HCWs of six national medical and research centers in Japan from October 2020 to March 2021. COVID-19-related discrimination was defined either when participants or their family members were badmouthed or when they felt discriminated against in some way. We used the Kessler Psychological Distress Scale (K6) to assess the presence of severe psychological distress (≥13 points). We used logistic regression models to examine the association between discrimination and psychological distress. We also identified job-related factors associated with discrimination.

**Results:** Of the participants, 484 (8.4%) reported COVID-19-related discrimination and 486 (8.5%) had severe psychological distress. HCWs who were female vs. male (odds ratio [OR]=1.41, 95% confidence interval [CI]=1.28-1.55), had high vs. low viral exposure (OR=2.31, 95%CI=1.81-2.93), and worked for more than 10 hours/day vs. <8 hours/day (OR=1.42, 95%CI=1.35-1.49) were more likely to have experienced COVID-19-related discrimination. The OR (95%CI) of severe psychological distress was 1.83 (1.29-2.59) among those who experienced discrimination. The analysis was stratified by sociodemographic and job-related factors and the associations trended in the same direction across subgroups.

**Conclusion:** Experience of COVID-19-related discrimination was associated with severe psychological distress among HCWs. During the pandemic, effective measures should be taken to prevent the development of negative mental health outcomes in HCWs who experience discrimination.

## Introduction

Since the emergence of ongoing coronavirus disease 2019 (COVID-19) pandemic, it has become a global health threat. Healthcare workers (HCWs), particularly those involved in COVID-19-related patient care were at a heightened risk of infection [1, 2]. For example, a meta-analysis, which including 28 studies from seven countries, reported that the percentage of HCWs who tested positive for COVID-19 was as high as 51.7% (95% confidence interval [CI] = 34.7–68.2) [1]. A prospective cohort study among 2,035,395 community individuals and 99,795 frontline HCWs reported that compared to the community individuals, frontline HCWs had a higher risk of infection (Hazard ratio [HR] = 11.6, 95% CI = 10.9–12.3) [2].

Since the beginning of the COVID-19 pandemic, the fear of transmission of the severe acute respiratory syndrome coronavirus 2 (SARS-CoV-2) from HCWs to the general population [3] provoked rapid stigma and discrimination towards HCWs, particularly against those involved in care of COVID-19 patients [4–6]. It was reported that HCWs faced discrimination in the form of verbal attacks and threats [3], avoidance from family and community members [7], avoidance from community members towards their family [8], and stigmatization [9]. Although fewer numbers of infections and death from COVID-19 have been reported in Japan compared to many other countries [10], a few studies reported that frontline HCWs and their family members have experienced discrimination [11, 12]. For instance, children of HCWs were refused access to kindergartens, school, and childcare facilities [11, 12].

Discrimination is an important determinant of negative mental health outcomes [13]. Given the concern regarding stigma and discrimination associated with COVID-19 during current pandemic, such experiences can lead to negative mental health consequences among the HCWs. For example, previous studies from Spain [14] and the Philippines [15] reported positive association between perceived discrimination and negative mental health outcomes such as depressive symptoms, psychological distress, and death thoughts. A survey in Japan among 4,386 HCWs reported that 19.1% felt avoided by their family members and friends [16]. However, there has been no study on the association between COVID-19-related discrimination and mental health among Japanese HCWs.

Thus, this study explored the factors associated with COVID-19-related discrimination and examined the association between experience of COVID-19-related discrimination and psychological distress among the staff of national medical research centers in Japan. We hypothesized that the experience of COVID-19-related discrimination could be positively associated with psychological distress among the HCWs. Furthermore, given that a certain group of HCWs (e.g., females and frontline workers) might be more susceptible to stigma and discrimination than other groups, we also hypothesized that the magnitude of association between discrimination and psychological distress may differ across subgroups in relation to socio-demographic and job-related factors.

## Methods

### Study design and participants

A multi-center collaborative study has been conducted among the staff members (mostly HCWs) of the six National Centers for Advanced Medical and Research in Japan to monitor the transmission of SARS-CoV-2. Each national center conducted a serological test and questionnaire survey at least once per year during the COVID-19 epidemic since 2020. Written informed consent was obtained from all the participants. After completing the opt-out process, the survey data were anonymized and submitted to the study committee for pooled analysis. The study design and procedure for data collection at each center were approved by the ethical committee of each center, while those of pooling study were approved by that of the National Center for Global Health and Medicine (NCGM) (approval number: NCGM-G-004233). For the current study, we used the data collected from the surveys conducted between October 2020 and March 2021 before the vaccination program at each center [17].

Of the 11,438 staff members invited for the survey, 5,919 participated (51.7% participation rate) (Figure 1). We requested all the eligible participants to complete questionnaire survey. After excluding participants without questionnaire data (n=120), with missing information on exposure (n=6), outcome (n=5), and selected covariates (described below) (n=81), 5,703 participants were included for the statistical analysis.

**Figure 1:**
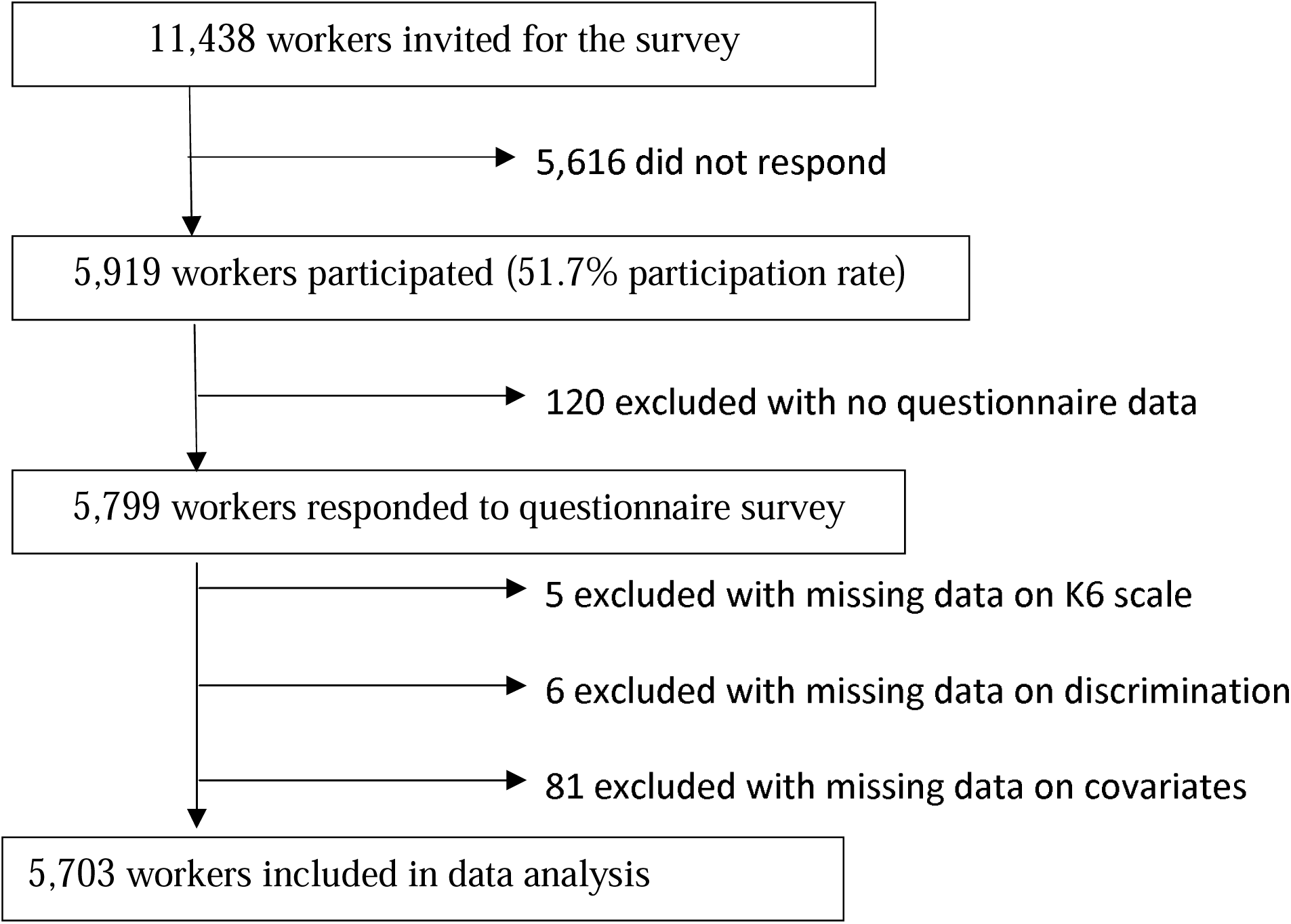
Flow diagram of study participants

### Measures

#### Psychological distress assessment (K6 scale)

The Japanese version of the Kessler Screening Scale for Psychological Distress (K6) scale was used to assess psychological distress [18]. It included six questions that rated participants’ frequency of how often they felt 1) nervous, 2) hopeless, 3) restless or fidgety, 4) so depressed that nothing could cheer them up, 5) that everything was an effort, and 6) worthless during the past 30 days. The responses options included five options, which ranged from “always” (score = 4) to “not at all” (score = 0) and the total score ranged from 0 to 24. Participants were judged to have severe psychological stress if the score was ≥13 points [19].

#### Discrimination

Regarding participants’ COVID-19-related experiences, we asked them if they agreed with the following two statements: “You and your family have been bad-mouthed” (yes or no) and “I felt that I was discriminated against in some way” (yes or no). If participants answered “yes” to at least one of the questions, they were considered to have experienced COVID-19-related discrimination. These questions were developed specifically for this survey based on a previous study conducted on COVID-19-related stigma among HCWs in Vietnam [20].

#### Covariates

We obtained participant’s information on the following covariates: sex, age, living arrangement, job category, COVID-19-related works, working hours,□smoking status, alcohol consumption, physical activity, sleep duration, height, weight, and comorbid□chronic□conditions.

□Job category included doctors, nurses, allied healthcare professionals, researchers, administrative and management staffs, and we merged researchers, administrative and management staffs into the category “non-clinical staffs”. Regarding engagement in COVID-19-related work, we asked the following two questions: “Have you ever engaged in the COVID-19 related work?” (yes or no) and “Did you engage in any work in which you were heavily exposed to the SARS-CoV-2?” (yes or no). We then defined the degree of possible exposure to SARS-CoV-2 at work and categorized the participants into three groups according to potential risk of infection: low (i.e. who did not engage in COVID-19 related work), moderate (i.e. engaged in COVID-19 related work, but without heavy exposure to the virus), and high (i.e. engaged in COVD-19 related work with heavy exposure to the virus).

Smoking status was categorized into three groups (never, former, and current smoker) based on participants’ responses on smoking conventional cigarettes and use of heated tobacco products (IQOS, glo, PULZE, WEECKE, etc.). Alcohol consumption was estimated based on the information on the consumption frequency and amount consumed in a day in “go” (*go* is a Japanese traditional unit equivalent to about 180ml). Leisure-time physical activity was measured in minute/week, based on one question about the weekly time spent on either indoor or outdoor physical activity.

Height and weight were used to calculate body mass index (BMI). We defined co-morbid condition if they had any one of the following chronic conditions: diabetes, hypertension, chronic obstructive pulmonary disease (COPD), heart disease, cerebrovascular disease, cancer, and other chronic diseases.

### Statistical analysis

First, we conducted multiple logistic regression analysis to investigate factors associated with COVID-19-related discrimination. We examined the associations in relation to age (<30, ≥30-<40, ≥40-<50, and ≥50 years), sex (male or female), living arrangement (living alone, and living with others), job categories (doctor, nurse, allied healthcare professional, and non-clinical staffs), degree of possible exposure to SARS-CoV-2 (low, moderate, and high), and working hours (≤_J8, 9 - 10, or ≥ 11 hours/day).

Then, we examined the association between discrimination and severe psychological distress via a multiple logistic regression analysis. The model was adjusted for the abovementioned variables as well as smoking status (never, former, and current smoker), alcohol consumption (<0.5, 0.5-<1, 1-<2, 2-<3, 3-<4, and ≥4 go/day), sleep duration (< 6; 6-<7 and ≥7 hours), comorbid conditions (yes or no), leisure-time physical activity (none, <1 hour/week, 1-<2 hours/week and ≥ 2 hours/ week) and BMI (< 18.5, 18.5 - <23, 23-<25, 25-<30, and ≥ 30kg/m^2^). We conducted stratified analyses by socio-demographic (age, sex and living arrangement) and occupation-related factors (job categories, degree of possible exposure to SARS-CoV-2 and working hours) to examine if the associations differ across the groups.

We reported OR and 95% CI for logistic regression and the level of significance was set at p*<* 0.05 (two-tailed). We used Stata version 15 (College Station, TX, USA) for all statistical analyses.

## Results

Table 1 presents the characteristics of study participants. Among 5,703 participants, 484 (8.4%) participants reported that they experienced a COVID-19-related discrimination (being bad-mouthed and/or experienced some sort COVID-19-related discrimination). In this study, 23.7% of the participants were below 30 years old, 70.2% were female and 67.5% were living with others. Regarding occupational background, 33.2% were nurses and more than half of the participants (59.7%) had lower degree of possible exposure to SARS-CoV-2.

**Table 1:** Characteristics of healthcare workers of six national medical research centers in Japan (N=5,703)

|  | Total<br>(n =5,703) | Discrimination |  |
| --- | --- | --- | --- |
|  |  | No<br>(n=5,218) | Yes<br>(n=484) |
| **Age, n (%)** |  |  |  |
| <30 | 1,353 (23.7) | 1,221 (90.2) | 132 (9.8) |
| 30-<40 | 1,464 (25.7) | 1,328 (90.7) | 136 (9.3) |
| 40-<50 | 1,568 (27.5) | 1,441 (91.9) | 127 (8.1) |
| ≥50 | 1,318 (23.7) | 1,229 (93.2) | 89 (6.8) |
| **Sex, n (%)** |  |  |  |
| Male | 1,702 (29.8) | 1,591 (93.5) | 111 (6.5) |
| Female | 4,001 (70.2) | 3,628 (90.7) | 373 (9.3) |
| **Living arrangement, n (%)** |  |  |  |
| Living alone | 1,851 (32.5) | 1,661 (89.7) | 190 (10.3) |
| Living with others | 3,852 (67.5) | 3,558 (92.4) | 294 (7.6) |
| **Job category, n (%)** |  |  |  |
| Doctors | 806 (14.1) | 745 (92.4) | 61 (7.6) |
| Nurses | 1,895 (33.2) | 1,664 (87.8) | 231 (12.2) |
| Allied healthcare professionals | 998 (17.6) | 922 (92.4) | 76 (7.6) |
| Non-clinical staffs | 2,004 (35.1) | 1,088 (94.2) | 63 (5.8) |
| **Degree of Possible exposure to SARS-CoV-2, n (%)** |  |  |  |
| Low | 3,407 (59.7) | 3,197 (93.8) | 210 (6.2) |
| Moderate | 1,181 (20.7) | 1,068 (90.4) | 113 (9.6) |
| High | 1,115 (19.6) | 954 (85.6) | 161 (14.4) |
| **Working hours, n (%)** |  |  |  |
| ≤ 8 hours/day | 3,027 (53.1) | 2,811 (92.9) | 216 (7.1) |
| 9-10 hours/day | 1,951 (34.2) | 1,758 (90.1) | 193 (9.9) |
| >10 hours/day | 725 (12.7) | 650 (89.7) | 75 (10.3) |
| **Smoking status, n (%)** |  |  |  |
| Never | 4,669 (81.9) | 4,280 (91.7) | 389 (8.3) |
| Former | 678 (11.9) | 625 (92.2) | 53 (7.8) |
| Current | 356 (6.2) | 314 (88.2) | 42 (11.8) |
| **Alcohol consumption, n (%)** |  |  |  |
| Don't drink | 2,155 (37.8) | 1,976 (91.7) | 179 (8.3) |
| < 1 go/day | 2,931 (51.4) | 2,682 (91.5) | 249 (8.5) |
| 1-<2 go/day | 487 (8.5) | 442 (90.8) | 45 (9.2) |
| ≥ 2 go/day | 130 (2.3) | 119 (91.5) | 11 (8.5) |
| **Leisure time physical activity, n (%)** |  |  |  |
| Not at all | 1,436 (25.2) | 1,323 (92.1) | 113 (7.9) |
| < 1 hour/week | 2,349 (41.2) | 2,165 (92.2) | 184 (7.8) |
| 1- < 2hours/week | 955 (16.7) | 864 (90.5) | 91 (9.5) |
| ≥ 2 hours/week | 963 (16.9) | 867 (90.0) | 96 (10.0) |
| **Sleep duration, n (%)** |  |  |  |
| < 6 hours/day | 2,692 (47.2) | 2,442 (90.7) | 250 (9.3) |
| 6-<7 hours/day | 2,052 (36.0) | 1,892 (92.2) | 160 (7.8) |
| ≥ 7 hours/day | 959 (16.8) | 885 (92.3) | 74 (7.7) |
| **BMI (kg/m<sup>2</sup>), n (%)** |  |  |  |
| < 18.5 | 631 (11.1) | 577 (91.4) | 54 (8.6) |
| 18.5-<23 | 3,387 (59.4) | 3,093 (91.3) | 294 (6.7) |
| 23-<25 | 832 (14.6) | 767 (92.2) | 65 (7.8) |
| 25-<30 | 697 (12.2) | 643 (92.3) | 54 (7.7) |
| ≥ 30 | 156 (2.7) | 139 (89.1) | 17 (10.9) |
| **Comorbid conditions, n (%)** |  |  |  |
| No | 4,350 (76.3) | 4,008 (92.1) | 342 (7.9) |
| Yes | 1,353 (23.7) | 1,211 (89.5) | 142 (10.5) |

Table 2 shows the results of multiple logistic regression analysis investigating the demographic and job-related factors associated with COVID-19-related discrimination. Female (OR = 1.41, 95%CI = 1.28–1.55), having moderate (OR = 1.50, 95%CI = 1.27–1.78), those with high exposure to SARS-CoV-2 (OR = 2.31, 95%CI = 1.81–2.93), and those working more than eight hours/day (OR = 1.26, 95%CI = 1.12–1.41) and more than ten hours/day (OR = 1.42, 95%CI = 1.35–1.49) were more likely to have the experience of COVID-19-related discrimination compared to male, those with low exposure to virus and those working short hours (≤ 8 hours/ day), respectively.

**Table 2:**
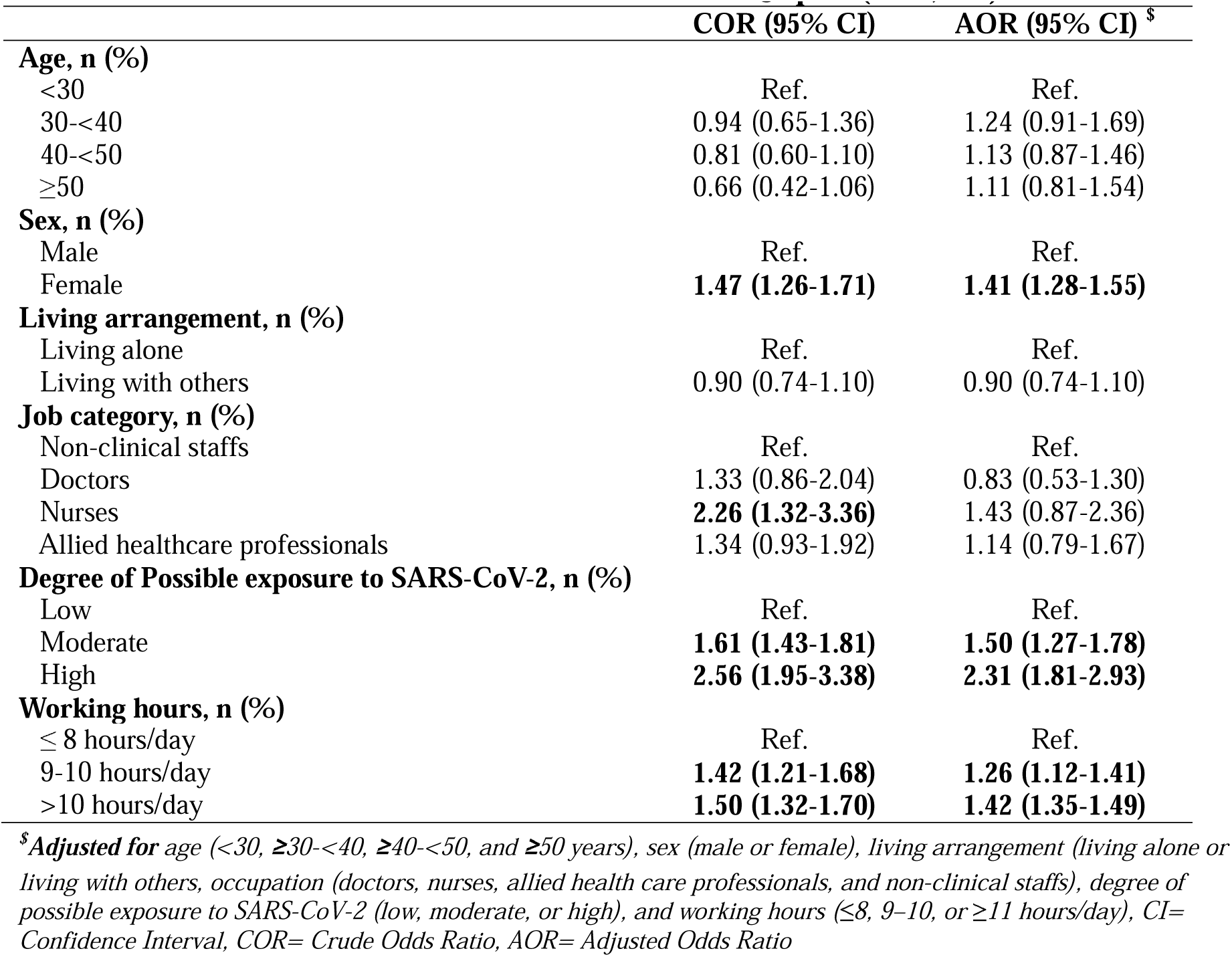
Factors associated with COVID-19-related discrimination among healthcare workers of six national medical research centers in Japan (N=5,703)

| | **COR (95% CI)** | **AOR (95% CI) <sup>\$</sup>** |
| --- | --- | --- |
| **Age, n (%)** |  |  |
| <30 | Ref. | Ref. |
| 30-<40 | 0.94 (0.65-1.36) | 1.24 (0.91-1.69) |
| 40-<50 | 0.81 (0.60-1.10) | 1.13 (0.87-1.46) |
| ≥50 | 0.66 (0.42-1.06) | 1.11 (0.81-1.54) |
| **Sex, n (%)** |  |  |
| Male | Ref. | Ref. |
| Female | **1.47 (1.26-1.71)** | **1.41 (1.28-1.55)** |
| **Living arrangement, n (%)** |  |  |
| Living alone | Ref. | Ref. |
| Living with others | 0.90 (0.74-1.10) | 0.90 (0.74-1.10) |
| **Job category, n (%)** |  |  |
| Non-clinical staffs | Ref. | Ref. |
| Doctors | 1.33 (0.86-2.04) | 0.83 (0.53-1.30) |
| Nurses | **2.26 (1.32-3.36)** | 1.43 (0.87-2.36) |
| Allied healthcare professionals | 1.34 (0.93-1.92) | 1.14 (0.79-1.67) |
| **Degree of Possible exposure to SARS-CoV-2, n (%)** |  |  |
| Low | Ref. | Ref. |
| Moderate | **1.61 (1.43-1.81)** | **1.50 (1.27-1.78)** |
| High | **2.56 (1.95-3.38)** | **2.31 (1.81-2.93)** |
| **Working hours, n (%)** |  |  |
| ≤ 8 hours/day | Ref. | Ref. |
| 9-10 hours/day | **1.42 (1.21-1.68)** | **1.26 (1.12-1.41)** |
| >10 hours/day | **1.50 (1.32-1.70)** | **1.42 (1.35-1.49)** |

<sup>\$</sup>*Adjusted for age (<30, ≥30-<40, ≥40-<50, and ≥50 years), sex (male or female), living arrangement (living alone or living with others), occupation (doctors, nurses, allied health care professionals, and non-clinical staffs), degree of possible exposure to SARS-CoV-2 (low, moderate, or high), and working hours (≤8, 9–10, or ≥11 hours/day), CI= Confidence Interval, COR= Crude Odds Ratio, AOR= Adjusted Odds Ratio*

Table 3 shows the results of multiple logistic regression analysis investigating the association between COVID-19-related discrimination and psychological distress. In this study, 486 (8.5%) had severe level of psychological distress. Compared with participants without experience of discrimination, OR (95% CI) of having psychological distress was 1.82 (95% CI = 1.30–2.55) among those who experienced discrimination. In stratified analyses by socio-demographic (age, sex and living arrangement) and occupation-related factors (job categories, degree of possible exposure to SARS-CoV-2 and working hours), the associations all trended in the same direction across the subgroups as indicated with a non-significant interaction.

**Table 3:** Overall association between discrimination and severe psychological distress and stratified association by socio-demographic and occupation related factors among healthcare workers (N=5,703)

| | Cases | Subjects | Discrimination | | P for interaction<sup>\$</sup> |
| --- | --- | --- | --- | --- | --- |
|  |  |  | No | Yes |  |
| **Overall** | 486 (8.5) | 5,217 (91.5) | 1.00 (Ref.) | **1.83 (1.29-2.59)** | — |
| **Age, n (%)** |  |  |  |  | 0.25 |
| <30 | 141 (10.4) | 1,212 (89.6) | 1.00 (Ref.) | 1.45 (0.67-3.10) |  |
| 30-<40 | 145 (9.9) | 1,319 (90.1) | 1.00 (Ref.) | **2.10 (1.74-2.53)** |  |
| 40-<50 | 128 (8.2) | 1,440 (91.8) | 1.00 (Ref.) | **2.15 (1.43-3.23)** |  |
| 50 | 72 (5.5) | 1,246 (94.5) | 1.00 (Ref.) | 1.91 (0.90-4.06) |  |
| **Sex, n (%)** |  |  |  |  | 0.78 |
| Male | 145 (8.5) | 1,557 (91.5) | 1.00 (Ref.) | 1.72 (0.78-3.77) |  |
| Female | 341 (8.5) | 3,660 (91.5) | 1.00 (Ref.) | **1.89 (1.45-2.46)** |  |
| **Living arrangement, n (%)** |  |  |  |  | 0.83 |
| Living alone | 200 (10.8) | 1,651 (89.2) | 1.00 (Ref.) | **1.68 (1.09-2.61)** |  |
| Living with others | 286 (7.4) | 3,566 (92.6) | 1.00 (Ref.) | **1.91 (1.20-3.05)** |  |
| **Job category, n (%)** |  |  |  |  | 0.28 |
| Doctors | 49 (6.1) | 757 (93.9) | 1.00 (Ref.) | **1.99 (1.15-3.46)** |  |
| Nurses | 201 (10.6) | 1,693 (89.4) | 1.00 (Ref.) | **1.83 (1.25-2.68)** |  |
| Allied healthcare professionals | 81 (8.1) | 917 (91.9) | 1.00 (Ref.) | **2.49 (1.62-3.84)** |  |
| Non-clinical staffs | 77 (9.0) | 776 (91.0) | 1.00 (Ref.) | 1.38 (0.70-2.68) |  |
| **Degree of Possible exposure to SARS-CoV-2, n (%)** |  |  |  |  | 0.24 |
| Low | 265 (7.8) | 3,142 (92.2) | 1.00 (Ref.) | 1.80 (0.95-3.41) |  |
| Moderate | 93 (7.8) | 1,088 (92.1) | 1.00 (Ref.) | 1.13 (0.61-2.10) |  |
| High | 128 (11.5) | 987 (88.5) | 1.00 (Ref.) | **2.58 (1.59-4.18)** |  |
| **Working hours, n (%)** |  |  |  |  | 0.68 |
| ≤ 8 hours/day | 220 (7.3) | 2,807 (92.7) | 1.00 (Ref.) | **2.02 (1.52-2.68)** |  |
| 9-10 hours/day | 175 (9.0) | 1,776 (91.0) | 1.00 (Ref.) | **1.72 (1.18-2.50)** |  |
| >10 hours/day | 91 (12.6) | 634 (87.4) | 1.00 (Ref.) | 2.10 (0.57-7.73) |  |

*Model was adjusted for age (<30, ≥30-<40, ≥40-<50, and ≥50 years), sex (male or female), occupation (doctors, nurses, allied health care professionals, and non-clinical staffs), living arrangement (living alone or living with others), degree of possible exposure to SARS-CoV-2 (low, moderate, or high), working hours (≤8, 9–10, or ≥11 hours/day), smoking (never, former, current), alcohol consumption (none, <1, 1–<2, or ≥2 go/day), sleep duration (<6, 6–<7, or ≥7 hours), comorbid conditions (yes or no), leisure-time physical activity (none, <1, 1–<2, ≥2 hours/week) and BMI (<18.5, 18.5–<23, 23–<25, 25–<30, or ≥30 kg/m<sup>2</sup>)*
*<sup>\$</sup>Derived from the joint test for the interaction between discrimination and covariates using “Contrast” command in Stata*

## Discussion

In this study, we identified female sex, the degree of possible exposure to SARS-CoV-2, and working hours as factors associated with COVID-19-related discrimination. Furthermore, the experience of COVID-19-related discrimination was positively associated with psychological distress. When we conducted the stratified analysis by socio-demographic and occupational factors, all the associations trended in the same direction.

We found that female HCWs were more likely to have experience of COVID-19-related discrimination compared with male HCWs. This finding was consistent with a previous study by Elhadi et al. [21] that reported higher stigmatization among female HCWs compared to male counterparts (36.1% vs. 28.2%). Staffs with higher exposure to SARS-CoV-2 being discriminated more is also comparable to a previous study by Yadav et al. [22] that showed higher perceived stigma among those working in high risk areas than those in low risk areas (73.7% vs. 67.4%). Furthermore, in this study, higher proportions of nurses (12.2%) experienced discrimination, followed by doctors and allied healthcare professions (7.6% each) and non-clinical staffs (5.6%). The results of multivariable analysis also suggested higher discrimination among the nurses (OR=1.43, 95% CI=0.87–2.36). This finding is in line with that of a previous study by Zandifar et al. [23] that reported higher discrimination among physicians and nurses than technicians. Regarding working hours, we found that those working longer hours tended to perceive higher discrimination. Healthcare workers who are working longer hours are known to be at higher risk for burnout [24], which could cause emotional exhaustion and have negative feelings about work [25] and might perceive higher sense of discrimination.

The significant association between COVID-19-related discrimination and psychological distress observed in this study was in line with that of a recent meta-analysis by Schubert et al., which reported the association between stigmatization from work-related COVID-19 exposure and depression (OR= 1.74; 95%CI = 1.29–2.36) and anxiety (OR = 1.75; 95% CI = 1.29–2.37) among HCWs during the COVID-19 pandemic [26]. These findings suggest that COVID-19-related discrimination could be harmful to mental health and should be addressed to ensure better mental health among frontline HCWs.

We expected that the extent of associations between COVID-19-related discrimination and psychological distress may significantly differ across the subgroups in relation with socio-demographic (age, sex and living arrangement) and job-related factors (job categories, degree of possible exposure to SARS-CoV-2 and working hours). However, we did not find any strong evidence of statistically significant interactions while the point estimates of the associations were higher among clinical staffs (doctors, nurses, and allied healthcare workers), HCWs with highest exposure to the virus compared with non-clinical staffs, and those with low exposure to the virus. A possible explanation for these findings could be that they were the ones involved in the treatment of COVID-19 patients. This could have made them more vulnerable to psychological distress associated with COVID-19-related discrimination they faced. Higher estimates of psychological distress among female HCWs in this study could be since they were more sensitive to COVID-19-related discrimination because of pre-existing discrimination and inequality against females [27].

The major strength of the present study includes large number of participants from six different national medical research centers in Japan. However, some limitations should be acknowledged. First, the information used in this study was self-reported, which could be subject to recall bias. Second, as the questionnaire included sensitive questions on mental health issues, responses could have been subject to social desirability bias. Third, we assessed psychological distress using a self-administered questionnaire via the K6 scale without administration by a psychiatrist. However, the scale has been validated in Japan [18]. Fourth, the questions for assessment of discrimination have not been validated. Fifth, because of the cross-sectional data, we do not know whether the associations are causal. Lastly, this study was conducted among those working in the healthcare and research centers, thus the findings may not be generalizable for other settings.

## Conclusion

This study provided evidence on the association between the experience of COVID-19-related discrimination and psychological distress among the HCWs from the six national healthcare centers in Japan. Our findings highlight the need of support for those who have suffered from mental health problems due to COVID-19-related discrimination.

## Data Availability

The data underlying this article cannot be shared publicly due to ethical restrictions and participant confidentiality concerns, but de-identified data are available from Dr. Mizoue (Department of Epidemiology and Prevention, Center for Clinical Sciences, National Center for Global Health and Medicine, Tokyo, Japan) to qualified researchers on reasonable request.

## References

1. Gholami M, Fawad I, Shadan S, et al (2021) COVID-19 and healthcare workers: A systematic review and meta-analysis. Int J Infect Dis IJID Off Publ Int Soc Infect Dis 104:335–346. https://doi.org/10.1016/j.ijid.2021.01.013

2. Nguyen LH, Drew DA, Graham MS, et al (2020) Risk of COVID-19 among front-line health-care workers and the general community: a prospective cohort study. Lancet Public Health 5:e475–e483. https://doi.org/10.1016/S2468-2667(20)30164-X

3. McKay D, Heisler M, Mishori R, et al (2020) Attacks against health-care personnel must stop, especially as the world fights COVID-19. The Lancet 395:1743–1745. https://doi.org/10.1016/S0140-6736(20)31191-0

4. World Health Organization (2020) Attacks on health care in the context of COVID-19. https://www.who.int/news-room/feature-stories/detail/attacks-on-health-care-in-the-context-of-covid-19. Accessed 20 Jan 2021

5. Devi S (2020) COVID-19 exacerbates violence against health workers. The Lancet 396:658. https://doi.org/10.1016/S0140-6736(20)31858-4

6. Vento S, Cainelli F, Vallone A (2020) Violence Against Healthcare Workers: A Worldwide Phenomenon With Serious Consequences. Front Public Health 8:570459. https://doi.org/10.3389/fpubh.2020.570459

7. Kirk AH, Chong S-L, Kam K-Q, et al (2021) Psychosocial impact of the COVID-19 pandemic on paediatric healthcareworkers. Ann Acad Med Singapore 50:203–211. https://doi.org/10.47102/annals-acadmedsg.2020527

8. Lau J, Tan DH-Y, Wong GJ, et al (2021) The impact of COVID-19 on private and public primary care physicians: A cross-sectional study. J Infect Public Health 14:285–289. https://doi.org/10.1016/j.jiph.2020.12.028

9. Greene T, Harju-Seppänen J, Adeniji M, et al (2021) Predictors and rates of PTSD, depression and anxiety in UK frontline health and social care workers during COVID-19. Eur J Psychotraumatology 12:1882781. https://doi.org/10.1080/20008198.2021.1882781

10. Iwasaki A, Grubaugh ND (2020) Why does Japan have so few cases of COVID 19? EMBO Mol Med 12:e12481. https://doi.org/10.15252/emmm.202012481

11. The Japan Times (2020) Front-line health care workers in Japan face discrimination over coronavirus. In: Jpn. Times. https://www.japantimes.co.jp/news/2020/05/29/national/health-care-workers-discrimination-coronavirus/. Accessed 10 May 2021

12. Jecker NS, Takahashi S (2021) Shaming and Stigmatizing Healthcare Workers in Japan During the COVID-19 Pandemic. Public Health Ethics phab003. https://doi.org/10.1093/phe/phab003

13. Pascoe EA, Richman LS (2009) Perceived Discrimination and Health: A Meta-Analytic Review. Psychol Bull 135:531–554. https://doi.org/10.1037/a0016059

14. Mediavilla R, Fernández-Jiménez E, Andreo J, et al (2021) Association between perceived discrimination and mental health outcomes among health workers during the initial COVID-19 outbreak. Rev Psiquiatr Salud Ment. https://doi.org/10.1016/j.rpsm.2021.06.001

15. Labrague LJ, De los Santos JAA, Fronda DC (2021) Perceived COVID 19 associated discrimination, mental health and professional turnover intention among frontline clinical nurses: The mediating role of resilience. Int J Ment Health Nurs 10.1111/inm.12920. https://doi.org/10.1111/inm.12920

16. Sahashi Y, Endo H, Sugimoto T, Nabeta T, Nishizaki K, Kikuchi A, et al. Worries and concerns among healthcare workers during the coronavirus 2019 pandemic: A web-based cross-sectional survey. Humanit Soc Sci Commun. 2021;8:41. https://doi.org/10.1057/s41599-021-00716-x

17. Yamamoto S, Tanaka A, Ohmagari N, et al (2021) Use of heat-not-burn tobacco products, moderate alcohol drinking, and anti-SARS-CoV-2 IgG antibody titers after BNT162b2 vaccination among Japanese healthcare workers. medRxiv

18. Furukawa TA, Kawakami N, Saitoh M, et al (2008) The performance of the Japanese version of the K6 and K10 in the World Mental Health Survey Japan. Int J Methods Psychiatr Res 17:152–158. https://doi.org/10.1002/mpr.257

19. Kessler RC, Barker PR, Colpe LJ, et al (2003) Screening for serious mental illness in the general population. Arch Gen Psychiatry 60:184–189. https://doi.org/10.1001/archpsyc.60.2.184

20. Do Duy C, Nong VM, Van AN, et al (2020) COVID 19 related stigma and its association with mental health of health care workers after quarantined in Vietnam. Psychiatry Clin Neurosci 10.1111/pcn.13120. https://doi.org/10.1111/pcn.13120

21. Elhadi M, Msherghi A, Elgzairi M, et al (2020) Burnout Syndrome Among Hospital Healthcare Workers During the COVID-19 Pandemic and Civil War: A Cross-Sectional Study. Front Psychiatry 11:579563. https://doi.org/10.3389/fpsyt.2020.579563

22. Yadav K, Laskar AR, Rasania SK (2020) A study on stigma and apprehensions related to COVID-19 among healthcare professionals in Delhi. Int J Community Med Public Health 7:4547–4553. https://doi.org/10.18203/2394-6040.ijcmph20204760

23. Zandifar A, Badrfam R, Mohammadian Khonsari N, et al (2020) Prevalence and Associated Factors of Posttraumatic Stress Symptoms and Stigma among Health Care Workers in Contact with COVID-19 Patients. Iran J Psychiatry 15:340–350. https://doi.org/10.18502/ijps.v15i4.4303

24. Lin R, Lin Y, Hsia Y, Kuo C (2021) Long working hours and burnout in health care workers: Non linear dose response relationship and the effect mediated by sleeping hours—A cross sectional study. J Occup Health 63: https://doi.org/10.1002/1348-9585.12228

25. Smith M, Segal J, Robinson L (2021) Burnout Prevention and Treatment -HelpGuide.org. In: https://www.helpguide.org. https://www.helpguide.org/articles/stress/burnout-prevention-and-recovery.htm. Accessed 13 Jun 2022

26. Schubert M, Ludwig J, Freiberg A, et al (2021) Stigmatization from Work-Related COVID-19 Exposure: A Systematic Review with Meta-Analysis. Int J Environ Res Public Health 18:6183. https://doi.org/10.3390/ijerph18126183

27. United Nations (2020) Women most affected by COVID-19, should participate in recovery efforts. https://www.ohchr.org/en/stories/2020/07/women-most-affected-covid-19-should-participate-recovery-efforts. Accessed 27 May 2022

